# Depression and Anxiety Symptoms among General Hospital Employees in need of Mental Health Treatment

**DOI:** 10.1101/2021.06.08.21258530

**Authors:** Nilson Silva, Anderson Sousa Martins da Silva, Lucas Pequeno Galvão, Julio Torales, Antonio Ventriglio, João Mauricio Castaldelli-Maia, Sergio Baldassin

## Abstract

**Background:** Depression and anxiety are common and disabling mental disorders worldwide. It has been described a high prevalence of these disorders among health professionals.

**Aim:** This study aimed to investigate the association between occupation and depressive/anxiety symptoms, education levels, among professionals from a Brazilian General Hospital in need of mental health treatment.

**Methods:** This is a longitudinal twelve-years retrospective study, involving health professionals. Socio-demographic data were collected as well as two standardized scales for depression and anxiety symptoms.

**Results:** Data from 506 employees needing a mental health intervention have been described: mean age was 34.6 years, 46.2% of them worked in the administrative sector, 35.0% were nursing assistants, 7.5% were nursing technicians, 6.7% were nurses, and 4.5% reported other occupations. According to the ICD-10 criteria, the rates of diagnosis of depressive disorders and anxiety disorders were 60.9% and 37%, respectively.

**Conclusions:** The rate of depression and anxiety is higher among health professionals than the general population. Thus, specific programs of prevention based on resilience, continuing education and health promotion are needed.

**Highlights:**

- Although 53,9% of studied individuals did not mention any kind of areas of difficulty in the workplace, 32.8% reported difficulties with coworkers or bosses.
- According to the ICD-10 criteria, diagnoses of depressive disorders (60.9%) and anxiety disorders (37%) were highly prevalent.
- Nursing technicians and nursing assistants reported higher somatic cluster BDI scores than other professionals of technical staff, but they were less inclined to receive a psychiatric diagnosis.

## Introduction

Depression disorders, burnout syndrome, anxiety disorders and suicides are common worldwide and frequent among health professionals (Dzau 2018; Edwards & Burnard, 2003; López-López, 2019; Melnick, 2020; Rotenstein, 2018; West, 2007; West, 2006), especially doctors and nurses, frequently due to their long training, work-related stress, personality characteristics as well as coping with illness and sick people in the daily life (Baldassin, 2003).

In 2005, our team set up a didactic ambulatory in São Paulo, Brazil, supporting hospital employees named *“Núcleo de Apoio aos Funcionários”* (NAF). Leaded by a professor of psychiatry of the medical school, it was created to offer psychiatric and psychological medical consultations. Senior medical undergraduate students of the FMABC Medical School are able to follow the psychiatrists and psychologists during the consultations. NAF consultations are standardized, and all patients are asked to respond in advance to The Spielberger State Anxiety Inventory (STAI-S) and Beck Depression Inventory (BDI). These data have been systematically recorded in a database.

Doctors and nurses report a relevant amount of distress (Foster, 2020). Also, women are two or three times more vulnerable: our study sample is composed predominantly by nurses and females. We aimed to provide a detailed report on health professionals mental health for promoting specific interventions and support.

### Research design and methods

This is a longitudinal twelve-years retrospective study based on a dataset collecting all psychiatric and psychological consults delivered for health professionals of a teaching hospital in São Paulo, Brazil with approximately 2.000 employees.

In this database, the following data have been collected: name, gender, degree of education, social status, marital status, date of birth, place of birth, place of residence, religion, pathway to care, professional area (technical or administrative activity), occupation, the main reason for searching help, clinical condition (mild, moderate or severe), years of profession, number of mental health professionals in the family, main areas of distress (bosses, coworkers, patients and none), if psychotherapeutic treatment was indicated, any previous psychiatric or psychological assessment/treatment, history of neuropsychiatric diseases or previous traumatic events during work.

Before the appointment, all patients filled out: 1] the Beck Depression Inventory (BDI) with 21 items(Gomes-Oliveira, 2012), divided in the affective cluster (BDI items: B1, B4, B10, B11, and B12), cognitive cluster (BDI items: B2, B3, B5, B6, B7, B8, B9, B13, B14, and B20), and somatic clusters (BDI items: B15, B16, B17, B18, B19, and B21) and 2] the State-Trait Anxiety Inventory STAI-S with twenty items (Andrade, 2001).

The following ranges of depressive symptoms have been considered for BDI: 0-9 - minimum or absent, 10-17 - mild, 18-29 - moderate, 30-63 - severe depression. The scoring for the STAI-S has been based on the following cut-offs: <33 - mild, 33-49 - moderate,>49 – high anxiety.

This study was approved by the research ethics committee under the number: 048/2010, on March 30, 2010, from Medical School of Health ABC University Center, Brazil.

Initially, data were analyzed descriptively. For the categorical variables, the absolute and relative frequencies were presented and for the continuous variables, summary measures (mean, quartiles, minimum, maximum and standard deviation) were used.

Bivariate analyses were performed with Chi-Square test, or alternatively in cases of small samples, Fisher’s exact test. In association, the standardized adjusted residue was used to identify local differences - cases with absolute values above 1.96 indicated evidence of (local) associations between the categories related to these cases.

The internal consistencies of BDI’s items (total and aspects) and STAI-S scales were evaluated using Cronbach’s Alpha (0.886 and 0.605, respectively).

The comparison of means between more than two groups were performed with the Variance Analysis (ANOVA). To detect differences in means, Duncan’s multiple comparisons were used. Once the differences in means in the ANOVA or Kruskal-Wallis test were detected, the differences location was discovered via multiple comparisons of Duncan or Dunn-Bonferroni, respectively maintaining a global significance level of 5%.

Finally, in order to evaluate the simultaneous effects of sex, age, race, religion, social status, education, occupation, area of difficulty, family member in mental treatment (explanatory variables) on BDI and STAI-S, multiple linear regression models were adjusted. Initially, all explanatory variables were included in the model and then the non-significant variables at 5% were excluded one by one in order of significance (backward method). Linear regression also presents as one of its assumptions the normality in the data, which was verified via the Kolmogorov-Smirnov’s test. For all statistical tests, a significance level of 5% was adopted. Statistical analyses were performed using Statistical Package for the Social Sciences 20.0.

## Results

In a twelve-years period, 4,168 accesses were performed at the didactic ambulatory in São Paulo for General Hospital employees, including 1,734 psychiatric medical appointments and 1,632 psychotherapy sessions.

Data from 506 patients were collected: mean age was 34.6 years (SD = 9.2 years), with a minimum age of 17 years old and a maximum age of 66 years old.

A predominance of women (85.8%) and professionals living with family (93.3%) have been recorded. In addition, about half of the professionals declared themselves as being Caucasian (55.0%) and catholic (45.5%).

Of 506 professionals, 234 (46.2%) worked in the administrative sector, 177 (35.0%) were nursing assistants, 38 (7.5%) were nursing technicians, 34 (6.7%) were nurses and 23 (4.5%) worked in other hospital technical staff, such as doctors, biomedicals, dentists, pharmacists, physiotherapists, phono-audiologists, nutritionists and pathologists.

A significant association has been found between occupation and education degree (p <0.001). Thus, it was observed that nurses and other professionals in the health technical staff had the highest percentages of individuals with higher education level (above 95%) when compared to nursing technicians and those in administrative positions (below 45.0%). The professionals with the highest percentage of higher education level were 33 (97.1%) nurses and 22 (95.7%) among the other technicians

There were no statistically significant differences between the incidence of mental disorders and factors such as social status, marital status, race or religion.

Table 1 shows that, out of 506 studied individuals, 262 (53.9%) did not indicate that the workplace was a source of distress, but 97 (20%) reported difficulties with coworkers, 62 (12.8%) with bosses and 53 (10.9%) with patients. The higher level of difficulties with patients was observed among nursing assistants (14.1%) and the lowest was observed among nurses (3.0%), which represents an observational finding with a non-significant Fisher’s exact descriptive test (p=0.610).

**Table 1.**
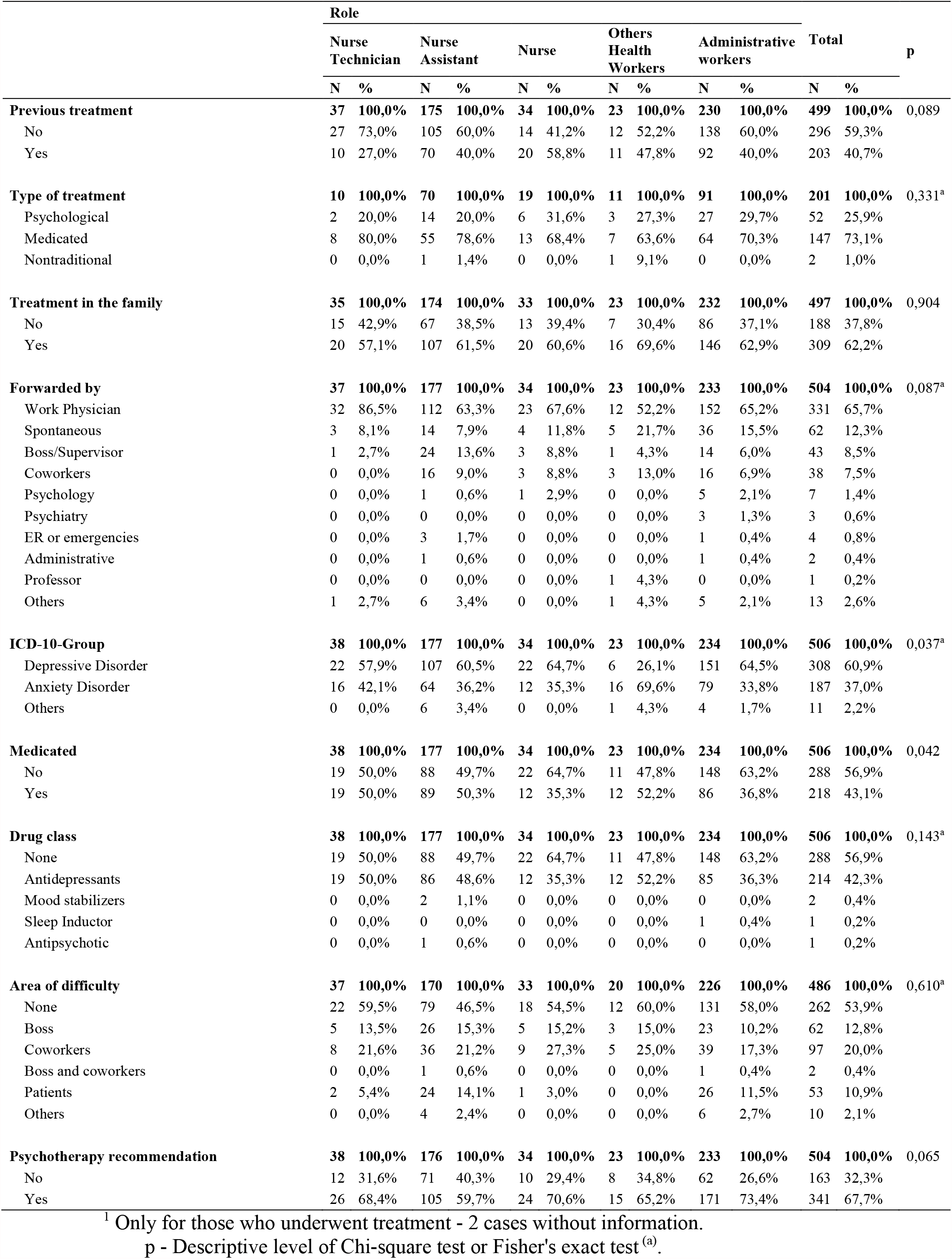
Distribution of professionals by clinical characteristics, according to occupation.

According to the ICD-10 (International Classification of Diseases-10th edition) criteria, diagnoses of depressive disorders (60.9%) and anxiety disorders (37%) were highly prevalent. Also, 69.6% of other technical staff professionals have been diagnosed with an anxiety disorder. (p=0.037).

Regarding the pharmacological treatment, when prescribed, antidepressants were largely employed in 43.1% (p=0.143) of professionals. For 341 of them (67.7%), psychotherapy has been indicated (p=0.065).

As shown in Table 1, an association between professional occupation and ICD-10 (p=0.037) as well as medication use (p=0.042) has been found. Thus, the other professionals of technical staff have shown a higher percentage of anxiety disorders (69.6%) when compared to other professionals which presented higher percentages of depressive disorders.

In general, it was observed that 40.7% received previous treatment (p=0,089), of which 73.1% pharmacological treatment (p=0,331). In addition, 62.2% reported a family history for neuropsychiatric treatments (p=0,904).

As shown in **Table 2** and **Figures 1** and **2,** there were differences between means of total BDI scores (p = 0.023), BDI affective cluster (p=0.019), and BDI somatic cluster (p=0.012) among patients with different healthcare-related occupations.

**Table 2.** Measures-summary of BDI (total and clusters) and STAI-S, according to occupation.

|  | Average | SD | Minimum | Maximal | 1<sup>st</sup><br>Quartile | Median | 3<sup>rd</sup><br>Quartile | N | p |
| --- | --- | --- | --- | --- | --- | --- | --- | --- | --- |
| **BDI** | **21,6** | **11,8** | **0,0** | **63,0** | **13,0** | **20,0** | **29,0** | **495** | **0,023** |
| Nurse's Technicians | 24,5<sup>(A)</sup> | 13,0 | 0,0 | 43,0 | 12,8 | 22,0 | 39,0 | 38 |  |
| Nurse's Assistants | 21,8<sup>(A)</sup> | 11,3 | 0,0 | 55,0 | 14,0 | 21,0 | 29,0 | 173 |  |
| Nurses | 21,9<sup>(A)</sup> | 12,7 | 0,0 | 49,0 | 12,0 | 18,0 | 30,5 | 33 |  |
| Others health technicians' staff | 15,1<sup>(B)</sup> | 10,2 | 1,0 | 43,0 | 9,8 | 13,0 | 17,3 | 22 |  |
| Administrative | 21,6<sup>(A)</sup> | 11,8 | 0,0 | 63,0 | 14,0 | 20,0 | 29,0 | 229 |  |
| **BDI-Affective** | **5,8** | **3,2** | **0,0** | **15,0** | **4,0** | **6,0** | **8,0** | **486** | **0,019** |
| Nurse's Technicians | 6,4<sup>(A)</sup> | 3,5 | 0,0 | 13,0 | 3,0 | 7,0 | 9,0 | 38 |  |
| Nurse's Assistants | 5,7<sup>(A)</sup> | 2,9 | 0,0 | 13,0 | 4,0 | 6,0 | 7,5 | 169 |  |
| Nurses | 6,0<sup>(A)</sup> | 3,4 | 0,0 | 13,0 | 4,0 | 6,0 | 8,0 | 34 |  |
| Others health technicians' staff | 3,8<sup>(B)</sup> | 2,9 | 0,0 | 12,0 | 1,0 | 4,0 | 5,0 | 23 |  |
| Administrative | 5,9<sup>(A)</sup> | 3,3 | 0,0 | 15,0 | 4,0 | 6,0 | 8,0 | 222 |  |
| **BDI-Cognitive** | **9,4** | **6,4** | **0,0** | **30,0** | **4,0** | **8,0** | **14,0** | **494** | **0,144** |
| Nurse's Technicians | 10,6 | 7,1 | 0,0 | 23,0 | 4,8 | 8,5 | 18,0 | 38 |  |
| Nurse's Assistants | 9,3 | 6,4 | 0,0 | 29,0 | 4,3 | 8,0 | 14,0 | 172 |  |
| Nurses | 9,1 | 6,7 | 0,0 | 22,0 | 3,0 | 8,0 | 15,3 | 34 |  |
| Others health technicians' staff | 6,6 | 5,7 | 0,0 | 20,0 | 3,0 | 4,0 | 9,0 | 23 |  |
| Administrative | 9,6 | 6,3 | 0,0 | 30,0 | 5,0 | 9,0 | 14,0 | 227 |  |
| **BDI-Somatic** | **6,3** | **3,8** | **0,0** | **18,0** | **3,0** | **6,0** | **9,0** | **496** | **0,012** |
| Nurse's Technicians | 7,2<sup>(A)</sup> | 4,3 | 0,0 | 18,0 | 4,0 | 7,5 | 9,3 | 38 |  |
| Nurse's Assistants | 6,7<sup>(A)</sup> | 3,8 | 0,0 | 16,0 | 4,0 | 6,0 | 9,0 | 173 |  |
| Nurses | 5,9 | 4,2 | 0,0 | 16,0 | 2,8 | 5,0 | 9,0 | 34 |  |
| Others health technicians' staff | 4,3<sup>(B)</sup> | 2,8 | 0,0 | 11,0 | 2,0 | 4,0 | 6,0 | 23 |  |
| Administrative | 6,0 | 3,7 | 0,0 | 18,0 | 3,0 | 6,0 | 8,0 | 228 |  |
| **STAI** | **44,9** | **6,7** | **25,0** | **65,0** | **41,0** | **45,0** | **49,0** | **492** | **0,942<sup>a</sup>** |
| Nurse's Technicians | 44,6 | 9,1 | 25,0 | 61,0 | 38,0 | 45,0 | 51,0 | 36 |  |
| Nurse's Assistants | 44,9 | 6,4 | 29,0 | 63,0 | 40,0 | 45,0 | 48,0 | 174 |  |
| Nurses | 44,7 | 6,6 | 30,0 | 59,0 | 40,0 | 45,0 | 48,5 | 33 |  |
| Others health technicians' staff | 46,1 | 5,2 | 38,0 | 60,0 | 43,0 | 45,0 | 47,5 | 21 |  |
| Administrative | 44,9 | 6,7 | 25,0 | 65,0 | 41,0 | 45,0 | 49,0 | 228 |  |

p - Descriptive level of ANOVA (a) or Kruskal-Wallis test
(A) and (B) present distinct means according to Dunn-Bonferroni comparisons

**Figure 1.**
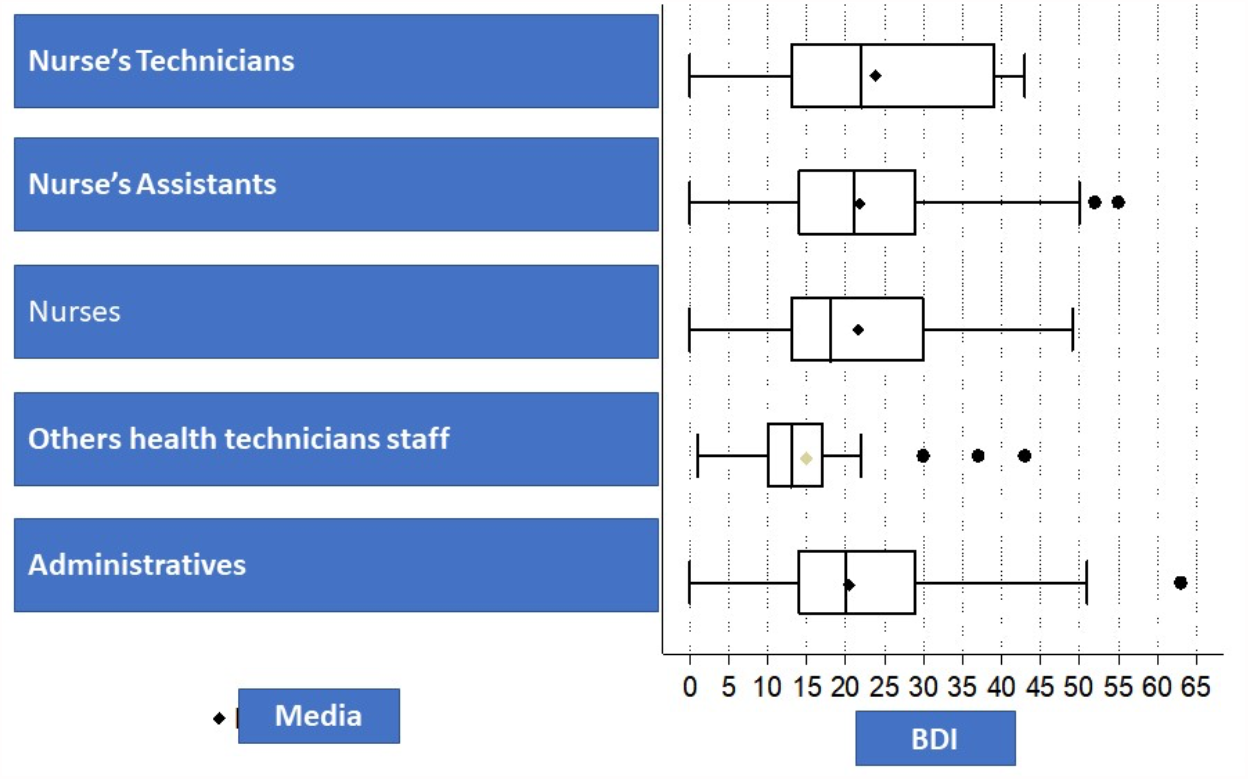
BDI Boxplot – total by professional role.

**Figure 2.**
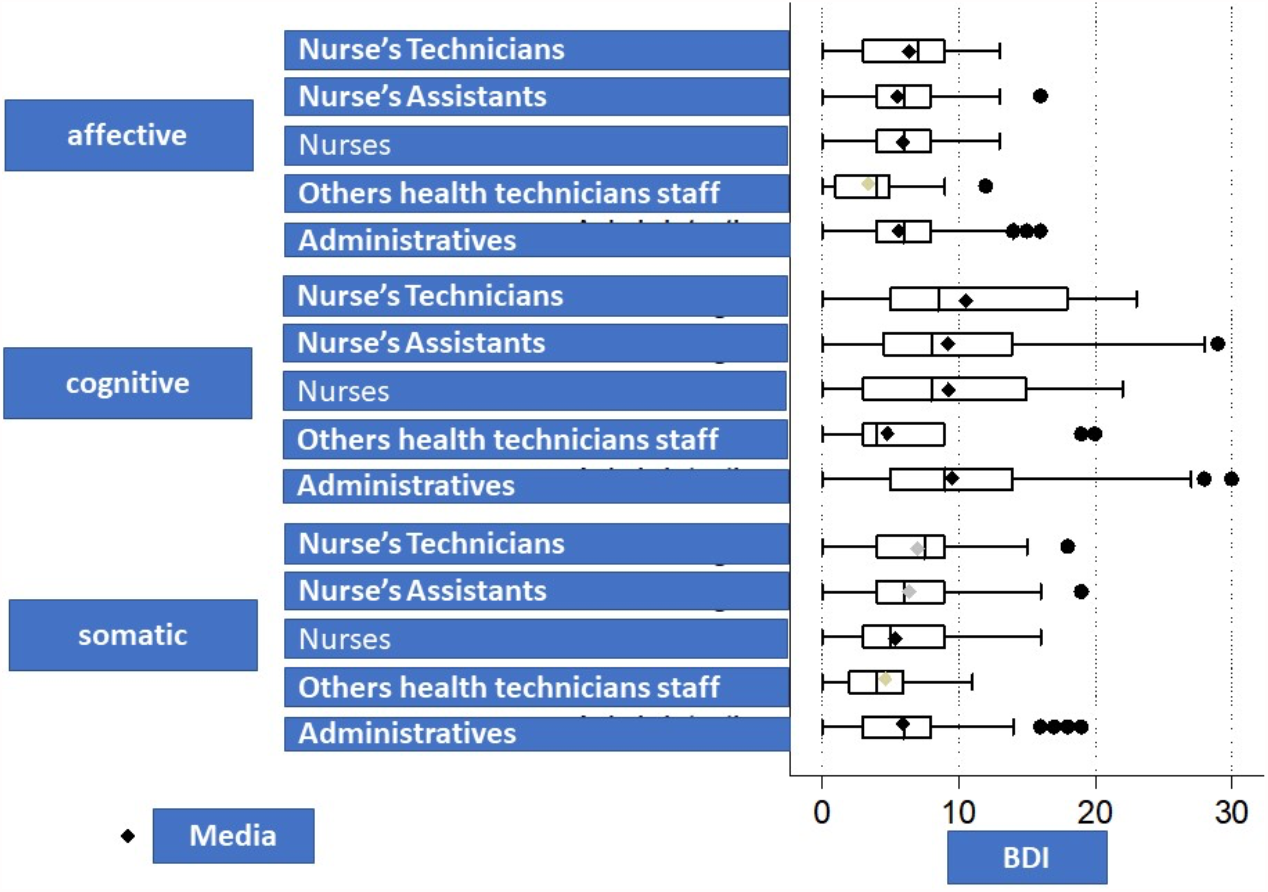
BDI clusters Boxplot by professional role.

Others professionals in the health technical staff presented lower means at total BDI score (p=0.023), BDI somatic cluster (p=0.012), and BDI affective cluster (p=0.019) than nurses, nursing assistants, nursing technicians and administrative staff, with the latter groups presenting similar means among themselves.

BDI cognitive cluster was not different among professional occupations (p=0.144) even if lower means of cognitive cluster have been found among administrative employees (not significantly).

Total BDI score (p=0.012), BDI somatic cluster (p=0.005), and STAI-S (p=0.012) were significantly different among professionals reporting areas of distress (**Table 3**). No associations were observed between STAI-S scores and occupation (p=0.276) and/or area of difficulty (p=0.085).

**Table 3.** BDI measures (total and clusters) and STAI-S by areas of difficulty.

| Area of difficulty | Average | SD | Minimum | Maximal | 1<sup>st</sup> Quartile | Median | 3<sup>rd</sup> Quartile | N | p |
| --- | --- | --- | --- | --- | --- | --- | --- | --- | --- |
| **BDI** |  |  |  |  |  |  |  |  | 0,012 |
| None | 20,4<sup>(B)</sup> | 12,2 | 0,0 | 55,0 | 12,0 | 18,0 | 28,0 | 259 |  |
| Boss | 23,7<sup>(B)</sup> | 10,4 | 0,0 | 49,0 | 18,0 | 22,0 | 31,0 | 60 |  |
| Coworkers | 22,5<sup>(B)</sup> | 11,4 | 0,0 | 63,0 | 13,0 | 22,0 | 29,0 | 95 |  |
| Boss and Coworkers | 10,5 | 2,1 | 9,0 | 12,0 | - | - | - | 2 |  |
| Patients | 22,9<sup>(B)</sup> | 11,6 | 1,0 | 63,0 | 16,0 | 21,0 | 31,0 | 51 |  |
| Others | 29,4<sup>(A)</sup> | 16,9 | 0,0 | 50,0 | 17,0 | 33,5 | 42,5 | 10 |  |
| **BDI-Affective** |  |  |  |  |  |  |  |  | 0,157<sup>a</sup> |
| None | 5,6 | 3,3 | 0,0 | 15,0 | 3,0 | 5,0 | 8,0 | 250 |  |
| Boss | 6,3 | 2,9 | 0,0 | 13,0 | 5,0 | 6,0 | 8,0 | 61 |  |
| Coworkers | 5,9 | 3,1 | 0,0 | 13,0 | 3,3 | 6,0 | 8,0 | 92 |  |
| Boss and Coworkers | 3,5 | 0,7 | 3,0 | 4,0 | - | - | - | 2 |  |
| Patients | 5,8 | 3,0 | 0,0 | 14,0 | 4,0 | 6,0 | 7,8 | 52 |  |
| Others | 7,8 | 4,1 | 0,0 | 13,0 | 4,8 | 9,5 | 11,0 | 10 |  |
| **BDI-Cognitive** |  |  |  |  |  |  |  |  | 0,063 |
| None | 9,1 | 6,6 | 0,0 | 30,0 | 4,0 | 8,0 | 14,0 | 257 |  |
| Boss | 10,3 | 5,8 | 0,0 | 24,0 | 6,0 | 10,0 | 15,0 | 60 |  |
| Coworkers | 9,8 | 6,5 | 0,0 | 28,0 | 5,0 | 9,0 | 13,0 | 95 |  |
| Boss and Coworkers | 0,5 | 0,7 | 0,0 | 1,0 | - | - | - | 2 |  |
| Patients | 9,1 | 5,8 | 0,0 | 19,0 | 5,0 | 8,0 | 15,0 | 51 |  |
| Others s | 12,5 | 8,6 | 0,0 | 25,0 | 5,3 | 12,0 | 19,8 | 10 |  |
| **BDI-Somatic** |  |  |  |  |  |  |  |  | 0,005 |
| None | 5,7<sup>(B)</sup> | 3,8 | 0,0 | 18,0 | 3,0 | 5,0 | 8,5 | 257 |  |
| Boss | 7,1<sup>(B)</sup> | 3,4 | 0,0 | 16,0 | 5,0 | 7,0 | 9,0 | 60 |  |
| Coworkers | 6,6<sup>(B)</sup> | 3,7 | 0,0 | 16,0 | 4,0 | 7,0 | 9,8 | 96 |  |
| Boss and Coworkers | 6,5 | 2,1 | 5,0 | 8,0 | - | - | - | 2 |  |
| Patients | 6,6<sup>(B)</sup> | 4,0 | 0,0 | 18,0 | 4,0 | 6,0 | 9,0 | 52 |  |
| Others | 9,3<sup>(A)</sup> | 5,3 | 0,0 | 14,0 | 5,5 | 12,0 | 14,0 | 10 |  |
| **STAI** |  |  |  |  |  |  |  |  | 0,012<sup>a</sup> |
| None | 44,0<sup>(A')</sup> | 6,7 | 25,0 | 65,0 | 40,0 | 44,0 | 48,0 | 254 |  |
| Boss | 46,8<sup>(B')</sup> | 6,4 | 34,0 | 63,0 | 42,0 | 47,0 | 52,0 | 61 |  |
| Coworkers | 45,5 | 6,6 | 29,0 | 61,0 | 41,3 | 45,0 | 50,0 | 96 |  |
| Boss and Coworkers | 43,0 | 5,7 | 39,0 | 47,0 | - | - | - | 2 |  |
| Patients | 46,6 | 6,5 | 31,0 | 62,0 | 41,0 | 46,0 | 50,0 | 51 |  |
| Others | 47,6 | 9,5 | 32,0 | 60,0 | 40,0 | 47,0 | 56,5 | 9 |  |

p - Descriptive level of ANOVA <sup>(a)</sup> or Kruskal-Wallis test
(A) and (B) present distinct means according to Dunn-Bonferroni comparisons
(A') and (B') present distinct means according to Duncan comparisons

After the application of a multiple linear regression model to total BDI (**Tables 4**), we found that professionals of technical staff recorded, on average, 7.04 points of depression symptoms less than the professionals working in the administrative area, although nursing technicians, nursing assistants and nurses presented similar levels to those in the administrative area (p=0.006). In addition, all professionals without family history of neuropsychiatric treatments recorded, on average, 3.64 points less in total BDI than those without this characteristic (p=0.001). Also, regarding the STAI-S (**Table 5**), we found that professionals reporting difficulties with bosses (p=0.018) or patients (p=0.039) registered, on average, 2.09 points of anxiety symptoms more than those who did not report any difficulty.

**Table 4.**
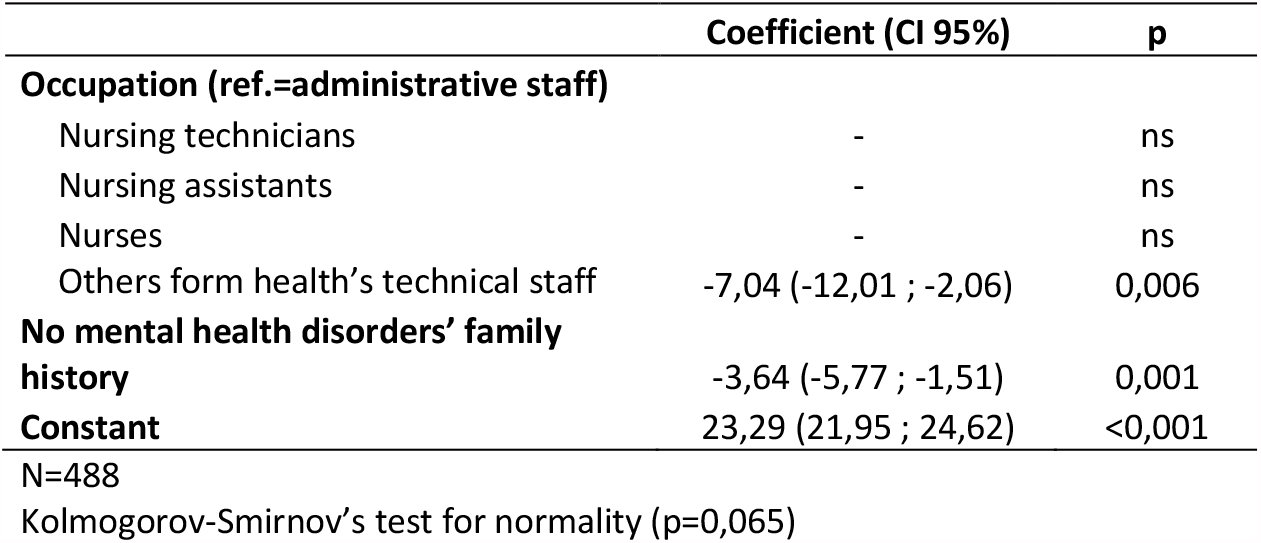
Multiple linear regression model’s estimates for total BDI.

**Table 5.**
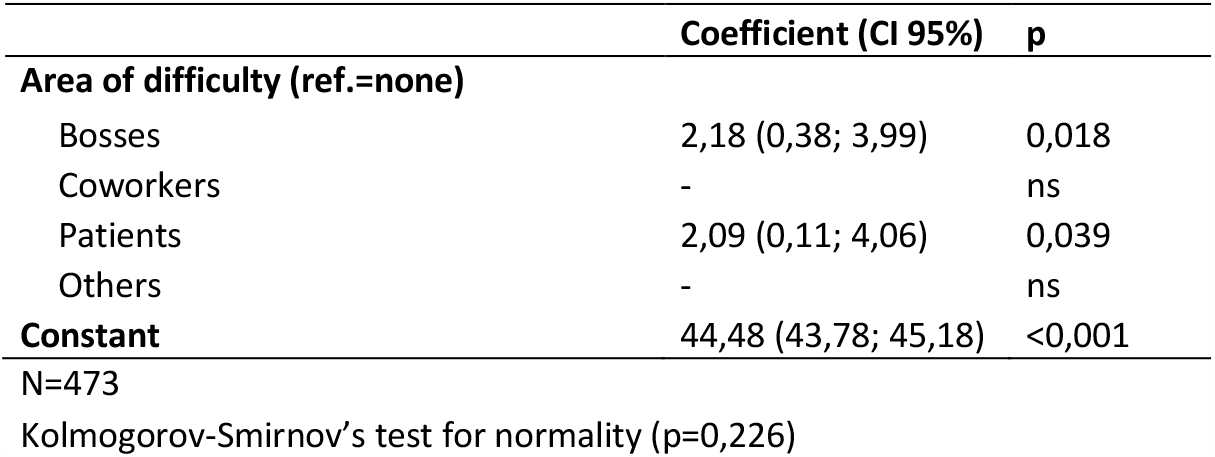
Multiple linear regression model’s estimates for STAI-S.

Analysis of internal consistency of BDI and STAI-S inventories showed a good internal consistency for BDI (total Cronbach’s Alpha = 0.886), and a moderated internal consistency for STAI-S (N=423), with a total Cronbach’s Alpha = 0.605.

## Discussion

Even before the SARS Cov-2 pandemic strike our fellow health professionals, most of professionals involved (65,7%) were referred by doctors of the Occupational Medicine Department and shortly after attending the didactic ambulatory, a high rate of drops-out have been observed.

It is of note that structured services, such as our didactic ambulatory, may improve the quality of treatments in mental health, such as for depressive and anxiety disorders (Eisenberg, 2007). In this study, for example, the fact that 46.2% of the patients came from the administrative area indicated that their managers should have provided specific interventions; also, nursing technicians (35%), reporting lower levels of education, might benefit from continuing education programs.

Although the majority of studied individuals (53,9%) did not mention any kind of areas of difficulty in the workplace, 32.8% reported difficulties with coworkers or bosses and could have received benefits from working resilience and social skills programs or, indirectly, from stimulating their bosses into changing their habits and routines in their administrative or technical approaches (Foster, 2018).

It is of interest that professionals with longer time of contact with patients, as nurses, nursing technicians and nursing assistants are frequently referred (49.2%) to the support service. This may be due to an exposure to higher level of stress and strategies aimed to improve their life quality may be developed (Delgado, 2020). Among these professionals, we found lower education levels, more females and nursing technicians.

Administrative staff employees also represented an important part (46,2%) of the patients but their characteristics and sources of difficulties are shown to be different. Total BDI scores (p=0,023) ranged as moderate depression (18-29 points in BDI), and along with BDI affective cluster (p=0,019) are higher among nurses, nursing technicians, nursing assistants and administrative staff than among other professionals of technical staff, reporting less contact with patients.

Nursing technicians and nursing assistants reported higher somatic cluster BDI scores (p=0,012) than other professionals of technical staff, but they were less inclined to receive a psychiatric diagnosis (Ross, 2009).

In the linear regression model, nursing technicians scored 4.93 (0.58;9.27, p=0.026) more points for depression at total BDI. Interventions aimed to improve physical and mental health among these professionals are recommended since they report exhausting job duties, more socioeconomic deficits, lower education and receive lower professional training.

They reported scores of seven points lower for depression than professionals from the administrative staff, as similarly described by Foster et al. 2020. Also, were more likely to be diagnosed with anxiety disorders, while other professionals reported higher rate of depressive diagnosis.

### Study Limitations

Limitations may include the involvement of one center only, even if data were systematically collected for twelve years and patients have been followed by the same professionals and with semi-structured academic standards.

### Implications

The existence of an independent academic nucleus to support hospital employees, in collaboration with the Occupational Medicine Department, has allowed the detection of mental health issues and associated factors among health care professionals. In addition, its activity has been also based on providing interventions for promoting knowledge about mental health, resilience with positive impact on health, quality of life of professionals and their families.

## Conclusion

The rate of depression and anxiety is higher among health professionals than the general population. Thus, specific programs, whenever possible connected with a medical school, of prevention based on resilience, delivering and promoting health education and involving senior doctors, hospital staff and patients to support the General Hospital employees, could be a great health goal, even after pandemics ends.

## Data Availability

All data is available in the article.

## Acknowledgements

To Prof. Milton Borelli and Dr. Angela Saragosa, who at the time directors of the public State Hospital Mario Covas, and Prof. Arthur Guerra de Andrade, Chair Professor of the Discipline of Psychiatry and Medical Psychology, facilitated the implementation of this service to support the hospital staff. To Dr. Eduardo Grecco, for the financial support of the performance of statistical work. To Dr. Ricardo Tenenbojm, Head of the Department of Occupational Medicine and nurses of the Department of Occupational Medicine: Glaucia Moda and Rafaela Silva, for the constant availability and gentle and understanding administrative interaction, and for the provision of physical and human resources, which supported us in the care and organization of this service. And of course, to all health professionals who participated in this study and to the medical students of FMABC who encouraged us to maintain this teaching outpatient clinic.

## ATTACHMENT

**Table I.1.** Kolmogorov-Smirnov’s test for data normality.

|  | Kolmogorov-Smirnov's test |  | N |
| --- | --- | --- | --- |
|  | z | p |  |
| Occupation |  |  |  |
| BDI | 1,72 | 0,005 | 495 |
| BDI - Affective | 1,48 | 0,025 | 486 |
| BDI - Cognitive | 2,02 | 0,001 | 494 |
| BDI - Somatic | 1,47 | 0,027 | 496 |
| STAI | 0,89 | 0,406 | 492 |
| Area of difficulty |  |  |  |
| BDI | 1,95 | 0,001 | 477 |
| BDI - Affective | 1,23 | 0,096 | 467 |
| BDI - Cognitive | 2,11 | <0,001 | 475 |
| BDI - Somatic | 1,80 | 0,003 | 477 |
| STAI | 0,71 | 0,697 | 473 |

**Table I.2.** Corrected item-total correlation and global Cronbach’s alpha (if an item is excluded from total BDI)

| Items | Item – Total Correlation | Cronbach's Alpha if the item was excluded |
| --- | --- | --- |
| **Global Cronbach's Alpha- 0,886** |  |  |
| Feeling sadness | 0,633 | 0,877 |
| Hopelessness | 0,555 | 0,879 |
| Feeling unsuccessful | 0,582 | 0,878 |
| Losing pleasure | 0,521 | 0,880 |
| Feeling guilty | 0,611 | 0,878 |
| Feeling punished | 0,439 | 0,884 |
| Disappointing yourself | 0,563 | 0,880 |
| Self-judging due to all things | 0,562 | 0,879 |
| Having suicidal thoughts | 0,503 | 0,881 |
| Crying | 0,446 | 0,882 |
| Getting angry | 0,395 | 0,884 |
| Disinterest for people | 0,556 | 0,879 |
| Being indecisive | 0,575 | 0,878 |
| Feeling old and ugly | 0,507 | 0,881 |
| Not being able to work | 0,563 | 0,879 |
| Losing sleep | 0,429 | 0,883 |
| Feeling too tired to act | 0,537 | 0,880 |
| Losing appetite | 0,484 | 0,881 |
| Losing weight | 0,169 | 0,890 |
| Concerning about physical health | 0,330 | 0,886 |
| Losing sexual interest | 0,476 | 0,882 |

N=420

On Table I.2, internal consistency values were shown (Global Cronbach’s alpha values and Cronbach’s Alpha values if an item was eliminated). Thus, it could be observed good internal consistency for total BDI (Cronbach’s Alpha = 0,886). Additionally, it was verified that not all item-total correlations presented values above 0,4, what indicated the presence of weak correlations between an item and the shaped score with all other scale’s items, excluding the participation of the item in question. However, it can be noticed that all items contribute favorably to internal consistency – the exclusion of an item doesn’t trigger a substantial raise in the value of Cronbach’s Alpha (Cronbach’s Alpha if the item was excluded).

**Table I.3.** Corrected item-total correlation and global Cronbach’s alpha (if an item is excluded from BDI’s clusters)

| Items | Item – Total Correlation | Cronbach's Alpha if the item was excluded |
| --- | --- | --- |
| **BDI-Affective (Global Cronbach's Alpha- 0,689)** |  |  |
| Feeling sadness | 0,528 | 0,602 |
| Crying | 0,421 | 0,649 |
| Losing pleasure | 0,466 | 0,629 |
| Disinterest for people | 0,435 | 0,643 |
| Getting angry | 0,370 | 0,669 |
| **BDI - Cognitive (Global Cronbach's Alpha - 0,827)** |  |  |
| Hopelessness | 0,548 | 0,808 |
| Feeling unsuccessful | 0,620 | 0,801 |
| Disappointing yourself | 0,599 | 0,806 |
| Feeling old and ugly | 0,482 | 0,816 |
| Feeling guilty | 0,614 | 0,803 |
| Feeling punished | 0,466 | 0,821 |
| Self-judging due to all things | 0,569 | 0,806 |
| Concerning about physical health | 0,321 | 0,830 |
| Being indecisive | 0,553 | 0,807 |
| Having suicidal thoughts | 0,492 | 0,816 |
| **BDI - Somatic (Global Cronbach's Alpha - 0,674)** |  |  |
| Not being able to work | 0,492 | 0,604 |
| Feeling too tired to act | 0,500 | 0,602 |
| Losing sleep | 0,382 | 0,641 |
| Losing appetite | 0,514 | 0,596 |
| Losing weight | 0,174 | 0,704 |
| Losing sexual interest | 0,398 | 0,637 |

As shown by table I.3, except for cognitive BDI (that showed a good internal consistency), all other clusters presented themselves with poor internal consistencies.

**Table I.4.** Corrected item-total correlation and global Cronbach’s alpha (if an item is excluded from STAI)

| Items | Correlation<br>Item -<br>Total | Cronbach's<br>Alpha if the<br>item was<br>excluded |
| --- | --- | --- |
| **Global Cronbach's Alpha - 0,605** |  |  |
| Feeling well | -0,055 | 0,619 |
| Getting easily tired | 0,269 | 0,585 |
| Wanting to cry | 0,328 | 0,577 |
| Wanting to be happy like others | 0,292 | 0,580 |
| Not being able to make quick decisions | 0,148 | 0,600 |
| Feeling rested | -0,003 | 0,621 |
| Feeling calm | -0,035 | 0,628 |
| Feeling difficulties building up | 0,309 | 0,578 |
| Worrying too much | 0,262 | 0,585 |
| Feeling happy | 0,172 | 0,597 |
| Letting yourself be affected | 0,344 | 0,573 |
| Not trusting yourself | 0,163 | 0,599 |
| Feeling safe | 0,110 | 0,607 |
| Avoiding to face crisis and problems | 0,231 | 0,589 |
| Feeling depressed | 0,329 | 0,576 |
| Feeling satisfied | 0,089 | 0,607 |
| Presence of irrelevant concerns | 0,251 | 0,587 |
| Taking yourself too seriously | 0,364 | 0,569 |
| Felling stable | 0,137 | 0,601 |
| Being tense | 0,372 | 0,570 |
| N=423 |  |  |

As shown by Table I.4, a poor internal consistency was verified for STAI (Cronbach’s Alpha = 0,605)

## References

Andrade, L., Gorenstein, C., Vieira Filho, A. H., Tung, T. C., & Artes, R. (2001). Psychometric properties of the Portuguese version of the State-Trait Anxiety Inventory applied to college students: Factor analysis and relation to the Beck Depression Inventory. Brazilian Journal of Medical and Biological Research, 34, 367–374.

Baldassin, S. (2003). Níveis, fontes e estratégias de enfrentamento de estresse psicológico entre estudantes de Medicina. Dissertação de Mestrado. Retrieved from dedalus.usp.br/F/FS3FBLYE64E3T19U4BL7LJJ3VVJRPSFK6KSNA95P1JETX8D6QL-05617?func=direct&doc%5Fnumber=001333882&pds_handle=GUEST.fromAGUIAhttp://dedalus.usp.br/F/FS3FBLYE64E3T19U4BL7LJJ3VVJRPSFK6KSNA95P1JETX8D6QL-05617?func=direct&doc%5Fnumber=001333882&pds_handle=GUEST

Delgado, C., Roche, M., Fethney, J., Foster, K. (2020) Workplace resilience and emotional labour of Australian mental health nurses: Results of a national survey. Int J Ment Health Nurs. 2020. 29(1),35–46. doi: 10.1111/inm.12598.

Dzau, V. J., Kirch, D. G., & Nasca, T. J. (2018). To Care Is Human - Collectively Confronting the Clinician-Burnout Crisis. N Engl J Med, 378(4), 312–314. doi:10.1056/NEJMp1715127

Edwards, D., & Burnard, P. (2003). A systematic review of stress and stress management interventions for mental health nurses. Journal of Advanced Nursing, 42(2), 169–200. doi:10.1046/j.1365-2648.2003.02600.x

Eisenberg, D., Golberstein, E., Gollust, S.E.(2007). Help-seeking and access to mental health care in a university student population. Med Care, 45(7), 594–601. doi: 10.1097/MLR.0b013e31803bb4c1. PMID: 17571007.

Foster, K., Cuzzillo, C., Furness, T.(2018) Strengthening mental health nurses’ resilience through a workplace resilience programme: A qualitative inquiry. J Psychiatr Ment Health Nurs. 25(5-6),338–348. doi: 10.1111/jpm.12467. Epub 2018 Jun 19. PMID: 29920873.

Foster, K., Roche, M., Giandinoto, J. A., & Furness, T. (2020). Workplace stressors, psychological well-being, resilience, and caring behaviours of mental health nurses: A descriptive correlational study. Int J Ment Health Nurs, 29(1), 56–68. doi:10.1111/inm.12610

Gomes-Oliveira, M. H., Gorenstein, C., Lotufo Neto, F., Andrade, L. H., & Wang, Y. P.. (2012). Validation of the Brazilian Portuguese version of the Beck Depression Inventory-II in a community sample. Brazilian Journal of Psychiatry, 34(4), 389–394. https://doi.org/10.1016/j.rbp.2012.03.005

López-López, I. M., Gómez-Urquiza, J. L., Cañadas, G. R., De la Fuente, E. I., Albendín-García, L., & Cañadas-De la Fuente, G. A. (2019). Prevalence of burnout in mental health nurses and related factors: a systematic review and meta-analysis. International Journal of Mental Health Nursing, 28(5), 1035–1044. doi:10.1111/inm.12606

Melnick, E. R., Dyrbye, L. N., Sinsky, C. A., Trockel, M., West, C. P., Nedelec, L., … Shanafelt, T. (2020). The Association Between Perceived Electronic Health Record Usability and Professional Burnout Among US Physicians. Mayo Clin Proc, 95(3), 476–487. doi:10.1016/j.mayocp.2019.09.024

Ross, C.A., & Goldner, E.M.(2009) Stigma, negative attitudes and discrimination towards mental illness within the nursing profession: a review of the literature. J Psychiatr Ment Health Nurs. 2009. 16(6), 558–67. doi: 10.1111/j.1365-2850.2009.01399.x. PMID: 19594679.

Rotenstein, L. S., Torre, M., Ramos, M. A., Rosales, R. C., Guille, C., Sen, S., & Mata, D. A. (2018). Prevalence of Burnout Among Physicians: A Systematic Review. JAMA, 320(11), 1131–1150. doi:10.1001/jama.2018.12777

West, C. P., Huntington, J. L., Huschka, M. M., Novotny, P. J., Sloan, J. A., Kolars, J. C., … Shanafelt, T. D. (2007). A prospective study of the relationship between medical knowledge and professionalism among internal medicine residents. Acad Med, 82(6), 587–592. doi:10.1097/ACM.0b013e3180555fc5

West, C. P., Huschka, M. M., Novotny, P. J., Sloan, J. A., Kolars, J. C., Habermann, T. M., & Shanafelt, T. D. (2006). Association of perceived medical errors with resident distress and empathy: a prospective longitudinal study. JAMA, 296(9), 1071–1078. doi:10.1001/jama.296.9.1071

